# Utilising novel Mendelian randomization approaches to investigate a potential causal effect of fetal growth on maternal cardiometabolic risk in the UK Biobank

**DOI:** 10.1101/2025.07.17.25331747

**Authors:** Alesha A Hatton, Caroline Brito Nunes, Christopher Flatley, Deborah A Lawlor, David M. Evans

## Abstract

Mothers of low birth weight infants are at increased risk of cardiovascular disease in later life. It is hypothesised this reflects a combination of 1) fetal factors influencing maternal vasculature via placental implantation, which also independently affect fetal growth, and 2) confounding by genetics and/or the environment whereby women with a pre-disposed risk of cardiometabolic dysfunction may have an impaired ability to adapt to the physiological stress of pregnancy. However, this does not preclude the possibility that there is a direct causal effect of fetal growth on maternal future cardiovascular health, with limited studies examining this alternative. Here, we applied two novel, complementary Mendelian randomization (MR) approaches to data from the UK Biobank that enabled us to investigate a potential causal effect of fetal growth (using birth weight as an available measure for fetal growth) on maternal cardiometabolic risk, even in the absence of offspring genotype information. A gene-by-environment interaction MR approach that stratified on maternal parity found no large differences in estimates of the causal effect of offspring birth weight on maternal cardiometabolic risk factors in parous and nulliparous females. Given that offspring birth weight cannot be causal for maternal cardiometabolic risk in nulliparous females, our results suggest that offspring birth weight is unlikely to causally affect maternal cardiometabolic risk factors. We also employed an offspring genotype by proxy MR design, which leveraged genotype and phenotype information from 45,546 spousal pairs. As fathers transmit half their alleles to their offspring, it follows that paternal genotype will proxy offspring genotype, which can in turn proxy offspring birth weight for MR analyses. This approach also found no strong evidence for a causal effect of offspring birth weight on maternal cardiometabolic risk factors although confidence intervals were wide. Our study serves as a blueprint for future investigations into the putative causal effects of offspring on their parents using MR.

## Background

The biological pathways that link pregnancy complications to the risk of later life cardiovascular disease (CVD) in women are not well understood. Extensive evidence supports the existence of common predisposing factors that contribute to both pregnancy complications, such as fetal growth restriction, and CVD risk (Fraser et al., 2018). Women with a predisposed risk of cardiometabolic dysfunction (either environmental or genetic) may have an impaired ability to adapt to the physiological stress of pregnancy, leading to an elevated risk of maternal and fetal pregnancy complications (Parikh et al., 2021). As such, pregnancy complications may serve as an early indication of heightened CVD risk trajectory, potentially preceding the clinical detection of traditional CVD risk factors. However, this does not preclude the possibility that there is a direct causal effect of fetal growth on maternal future cardiovascular health, with limited studies examining this alternative (Rich-Edwards et al., 2014).

Fetal growth is regulated by a complex interplay of maternal, fetal and placental factors. This process depends on proper placental implantation and sufficient blood flow through the maternal uterine arteries, which supply oxygen and nutrients to the fetus. Evidence suggests that mechanisms driving fetal growth influence maternal physiology during pregnancy, potentially enhancing blood supply and nutrient transfer across the placenta. Within the fetal genome, paternally transmitted birth-weight increasing alleles have been associated with reduced gestational duration and increased maternal systolic blood pressure during pregnancy (Chen et al., 2020). Similarly, fetal genetic predisposition to a higher placental weight raises the risk of maternal preeclampsia and shortens gestational duration (Beaumont et al., 2023). Further, paternally-transmitted fetal alleles have been associated with increased late pregnancy maternal glucose concentrations (Petry et al., 2017; Petry et al., 2011), blood pressure (Petry et al., 2016) and gestational hypertension (Petry et al., 2016). While most of maternal cardiovascular adaptations during pregnancy resolve in the postpartum period, it remains unclear whether mechanisms that influence fetal growth have lasting metabolic and vascular effects on the mother.

Much of the research associating fetal growth with maternal CVD risk later in life has been derived from observational studies through the linkage of large birth registries with hospitalization and mortality statistics (Haug et al., 2021; Morken et al., 2018; Shaikh et al., 2018; Shaikh et al., 2020; Smith et al., 2000; Smith et al., 2005). While these have produced consistent associations of fetal growth with CVD risk, such observational studies may be limited in their ability to dissect the complex interplay between maternal and offspring influences. Since mothers transmit 50% of their genes to their offspring (and given that genetic pleiotropy is likely to be pervasive in the human genome), genetic confounding is likely to explain at least some of the observational correlation between maternal and offspring phenotypes (Bond et al., 2020; Shaikh et al., 2018; Shaikh et al., 2020). Owing to the advent of large genetic datasets, methods for improving our understanding of whether pregnancy complications are causally related to later maternal health are now able to go beyond conventional observational approaches.

Mendelian randomization (MR) is an epidemiological technique that uses genetic variants as instrumental variables to estimate the causal effect of an exposure on medically relevant outcomes (Smith & Ebrahim, 2003). Mendel’s Laws of Segregation and Independent Assortment ensure (with certain important exceptions (Lawlor et al., 2008)) that genetic variants are randomized with respect to potential genetic and environmental confounders. This means that MR studies are robust to some of the issues faced by traditional observational epidemiological approaches such as confounding and reverse causality, though have different sources of bias (Brumpton et al., 2020). In theory it is possible to use offspring genetic variants to proxy offspring birth weight and then use MR to estimate the causal effects of offspring birth weight on maternal cardiometabolic risk (Chen et al., 2020).

However, maternal and offspring genotypes are correlated. Consequently, any association between the offspring genotype and maternal cardiometabolic risk, when no adjustment has been made for maternal genotype, may reflect an effect of maternal genotype on maternal outcomes. Therefore, to avoid confounding through the maternal genome, it is necessary to condition the analysis on maternal genotype. A similar approach can be employed (which avoids genetic confounding through the maternal genome) that makes use of paternally transmitted alleles, either inferred from phased mother-offspring pairs or from genotyped parent-offspring trios to proxy offspring phenotype. Chen et al. used this method to demonstrate offspring birth weight scores constructed from paternally transmitted alleles were associated with reduced gestational duration and increased maternal systolic blood pressure during pregnancy (Chen et al., 2020). Whilst good in theory, broader application of these approaches at scale is hampered by the paucity of genotyped mother-offspring pairs (Evans et al., 2019) meaning that such analyses may have low power to resolve causal effects.

In this study we investigated whether there is a direct causal effect of fetal growth on maternal future cardiovascular health. To do this, we leveraged two complementary novel MR approaches that can be employed in the absence of offspring genetic information. First, we employed a variant of the MR gene-by-environment interaction (MR GxE) approach (Davey Smith, 2011; Spiller et al., 2019), using parity in females as the environmental variable on which to stratify analyses. If offspring birth weight is causal for maternal cardiometabolic risk factors, it follows that any causal effect should be observed in parous females, but not in nulliparous females. The difference in causal effect estimates between the two groups should also provide an estimate of the true causal effect corrected for any latent horizontal pleiotropy. Second, we utilized fathers’ genotypes from the estimated 45,546 spousal pairs in the UK Biobank (Horwitz et al., 2023; Tenesa et al., 2016) to proxy offspring birth weight. As fathers transmit half their alleles to their offspring, it follows that paternal genotype at known birth weight associated loci will proxy offspring genotype, which in turn will proxy offspring birth weight. The advantage is that paternal genotype should be unrelated to maternal genotype (in the absence of assortative mating for birth weight and related traits). This makes the approach robust to genetic confounding through the maternal genome (and of course assuming paternal genotype does not directly affect maternal outcome), and leverages previously unused information on the tens of thousands of spousal pairs in the UK Biobank. Jointly these approaches allowed us to interrogate the potential presence of a causal effect of fetal growth on maternal cardiometabolic risk.

## Methods

### UK Biobank

The UK Biobank (UKB) is a large prospective population-based cohort of over 500,000 participants, aged between 37-73 years (54% female) at baseline (Sudlow et al., 2015). UKB has ethical approval from the North West Multi-Centre Research Ethics Committee, which covers the UK, and all participants provided written informed consent. The cohort has been described in detail previously (Bycroft et al., 2018; Sudlow et al., 2015) and contains a broad range of health-related information and genome-wide genetic data. The full data release in the UKB contains a cohort of 502,511 successfully genotyped individuals. Pre-imputation quality control, phasing and imputation are described in Bycroft et al., (Bycroft et al., 2018). In addition to the quality control metrics performed centrally by the UKB, we excluded individuals with withdrawn consent, sex-mismatch (derived by comparing genetic sex and reported sex), sex chromosome aneuploidy, and high heterozygosity or missingness (n= 500,639). Individuals were restricted to those of European descent using a K-means (K = 4) clustering approach based on the first four genetically determined principal components (n=477,098). We identified all pairs of related individuals using KING software (107,119 pairs of relateds). Relatedness filtering was performed where one individual from every related pair was excluded, resulting in 211,557 female participants and 45,546 spousal pairs.

### Phenotypic information

We used baseline data measured (or self -reported) at the UKB initial assessment centre. Parity was determined for female individuals using their self-reported number of offspring and for males using their self-reported number of children fathered. Females also self-reported their age at first and last birth. Females were asked to report the birth weight of their first child to the nearest pound (that we converted to kg). Data on gestational duration were not available; however, in order to exclude likely pre- and post-term births, participants with birth weight values <2.5kg (n=13,341) or >4.5kg (n=2,703) were excluded. 156 measurements were also excluded because the multiple reports of offspring birth weight differed by >1 kg. Following QC, there were 148,080 females with the birth weight of their first child reported. Participants were also asked to report their own birth weight with data for 180,469 individuals (excluding 5,905 individuals who were part of multiple births, 46 where the multiple reports of offspring birth weight differed by >1 kg, 36,198 individuals who reported their own birth weight to be <2.5 kg (n= 18,271) or >4.5 kg (n= 10,807)).

Maternal cardiometabolic risk outcomes explored were systolic blood pressure (SBP), diastolic blood pressure (DBP), glucose (Gl), total cholesterol (TC), high density lipoprotein cholesterol (HDL), low density lipoprotein cholesterol (LDL), triglycerides (TG), and body mass index (BMI). SBP and DBP (mmHg) were measured using a digital monitor, or a manual sphygmomanometer if the digital monitor could not be employed. Participants provided a non-fasting blood sample, which was assayed for a panel of biomarkers using standard laboratory procedures (http://Biobank.ndph.ox.ac.uk/showcase/refer.cgi?id=5636). Non-fasting serum biochemical measurements including Gl, TC, HDL, LDL and TG. BMI (kg/m2) was constructed from measured standing height and weight. Variables that were not normally distributed (TG, BMI, and Gl) were first natural log transformed. All outcome variables were then normalised (mean zero, variance 1). Values more than 4 standard deviations from the mean for each outcome were removed.

Covaraites used for analysis included age at assessment, assessment centre, genotyping batch, the first 20 genetic PCs. Additional potential confounders were smoking (current, previous or never), alcohol intake frequency (converted to units per month), educational qualifications as a proxy for years of education and residential area deprivation as measured using the Townsend deprivation index. Information on which of the 22 centres in Scotland, England and Wales where assessments were undertaken was provided. Years of education was constructed by mapping each educational qualification to the equivalent years of education using the International Standard Classification for Education (ISCED) coding as performed previously (Lee et al., 2018). Descriptive statistics are presented in Table S1.

### Identifying spouse pair subset

Spousal information is not explicitly available in the UK Biobank. Potential spouses were therefore identified using phenotypic data based on the definition in Tenesa et al. (Tenesa et al., 2016) in addition to six further criteria (Table S2). Firstly, potential spousal pairs were identified using the returned dataset with potential couples provided by Tenesa et al., (i.e., where colocation information along with sex, year/month of birth, how people in the household are related to the participant, number of individuals in household and Father’s/Mother’s ages and ages at death were used to identify spousal pairs). Pairs were removed if both individuals were related (IBD>0.08). Putative couples were then matched based on their responses to five additional household variables. These included their responses to questions on the number of vehicles, whether they owned/rented the accommodation they lived in, type of accommodation they lived in, household income and Townsend Deprivation Index. Participants were required to provide identical responses for these variables, with the exception of household income, where pairs were still matched if their reported income differed by one category or if they had missing income information. Spousal pairs were then matched on the same number of reported offspring. As only female participants in UKB report the birth weight of their first offspring, matching phenotypic data between spousal pairs allowed us to also report the offspring birth weight for male spouses. This resulted in 45,546 spousal pairs with offspring birth weight recorded. As a validation step, we cross-checked with parent-offspring trios identified based on genetic information and found the criteria used in the matching of spousal pairs to be largely concordant in this subset, suggesting the spousal pairs derived in this study are genuine (Supplementary Material 1).

### Multiple Testing Correction Threshold

Principal component analysis across the 8 outcomes suggested an approximate multiple-testing corrected P-value threshold of P < 0.0125 for statistical significance for all analyses conducted (see details in Supplementary Material 2).

### Traditional observational epidemiological analyses

Phenotypic associations between offspring birth weight and maternal cardiometabolic risk factors later in life were assessed in 151,478 females in the UKB using multivariable linear regression. All models were adjusted for standard covariates and potential confounders (as described above). We also repeated the analysis in the subset of females with a matched spouse (n=45,546) as differences in the phenotypic association in this subset may indicate selection bias. Lastly, using the offspring birth weight reported by the female spouse, we were also able to assess the relationship between offspring birth weight and paternal cardiometabolic risk factors in the spousal pairs (n=45,546). All linear regression analyses were conducted using R version 4.3.0.

### Mendelian Randomization Analyses

Individual-level and two-sample MR analyses were performed to examine the relationship between offspring birth weight and maternal cardiometabolic risk factors in later life. Power calculations for these analyses are presented in Supplementary Material 3. For all analyses we included age at assessment as a negative control outcome.

#### The selection of genetic instruments

Two strategies were used to proxy offspring birth weight in the MR analyses. First, we used a paternal PGS for (father’s) own birth weight, comprised of genetic variants that have been associated with own birth weight. Here we assumed that paternal genetic variants for own birth weight influence offspring birth weight through transmission of alleles from father to offspring. Second, we used a maternal PGS for offspring birth weight which was comprised of maternal genetic variants that have been associated with offsprings birth weight. We subsequently describe the selection of genetic variants and the construction of these two PGS.

##### Identification of genetic variants for own and offspring birth weight

SNPs associated with own (n=423,683) and offspring birth weight (n=270,002) were identified from the GWAS meta-analysis of the Early Growth Genetics (EGG) Consortium, UKB and deCODE (Juliusdottir et al., 2021) cohorts. Due to the correlation between the maternal and fetal genomes, Hwang et al. (Hwang et al., 2024) partitioned genetic effects on birth weight into maternal and fetal contributions. Specifically, 381 independent SNPs were identified to be associated with birth weight, of which 369 were from the GWAS meta-analysis (P-value < 3.3 x 10^-9^) and an additional 12 loci from a two-degree of freedom test combining GWAS of own and offspring’s birth weight (P-value < 6.6 x 10^-9^). SNPs with significant fetal genetic effects (P-value < 0.05) influence birth weight directly through the fetal genome, while those with significant maternal genetic effects (P-value < 0.05) are thought to act indirectly on offspring birth weight through the maternal genome. These SNPs were extracted from UKB imputed genotype data in dosage format using Plink (v2.0) (Chang et al., 2015) (excluding four SNPs on the X chromosome).

##### Paternal PGS for own birth weight

We used a paternal PGS for (father’s) own birth weight, as a proxy for offspring PGS at the same loci, which in turn proxied offspring birth weight. In other words, we assume that paternal genetic variants for own birth weight influence offspring birth weight through transmission of alleles from father to offspring. Paternal PGS were constructed from combinations of 381 independent SNPs known to be associated with own and/or offspring birth weight to identify the combination of genetic variants most strongly associated with offspring birth weight. PGS were calculated as the sum of all 381 lead birth weight associated SNPs available in the UKB (F1), 287 autosomal SNPs that showed evidence of a fetal effect (F2) and 205 autosomal SNPs that showed only significant (P-value < 0.05) fetal effects on birth weight (F3) as described above (Table S3). Dosages were coded so that increases reflected alleles associated with increased own birth weight. Missing genotypes were imputed as two times the frequency of the own birth weight increasing allele. We constructed both weighted and unweighted scores. We employed this approach owing to sample overlap between the discovery GWAS and the analysis sample. For unweighted scores, the weight for each allele was set as one. For weighted scores, weights were obtained from the estimated fetal effects reported in Hwang et al. (Hwang et al., 2024). For all approaches, scores were calculated in the spousal, parous fathers where the mother had reported offspring’s birth weight (n= 45,546) using the profile scoring routine in the Plink (v2.0). For each individual, scores were calculated as the sum of birth weight increasing alleles, across all SNPs multiplied by the corresponding weight.

##### Maternal PGS for offspring birth weight

We employed a similar approach to proxy offspring birth weight, again constructing PGS from combinations of the 381 independent SNPs associated with own or offspring birth weight to identify the combination of genetic variants most strongly associated with offspring birth weight. PGS were calculated as the sum of all 381 autosomal lead birth weight associated SNPs available in the UKB (M1), 176 autosomal SNPs that showed significant evidence of a maternal effect (M2) and 94 autosomal SNPs that showed evidence of only maternal effects on offspring birth weight (M3) (Table S3). Dosages were coded so that increasing dosage reflected alleles associated with increased offspring birth weight. Missing genotypes were imputed as two times the frequency of the offspring birth weight increasing allele. Again, we constructed both weighted and unweighted scores. For unweighted scores, the weight for each allele was set as one. For weighted scores, weights were obtained using maternal effects reported in Hwang et al. For all approaches, scores were calculated in 171,031 parous and 40,239 nulliparous females in the UKB using the profile scoring routine in the Plink (v2.0).

#### Gene by Environment Mendelian randomization

We utilised the GxE MR framework (Chen et al., 2008; Davey Smith, 2011) to investigate evidence of a causal association between offspring birth weight and maternal cardiometabolic risk factors in the UKB (Figure S1). Here, we tested whether maternal PGS that are indirectly associated with offspring birth weight (i.e. maternal SNPs that have maternal indirect effects on offspring birth weight) were also associated with maternal cardiometabolic risk. While the PGS used primarily capture indirect maternal effects on offspring birth weight, the M1 and M2 PGS also include SNPs with fetal effects on own birth weight. These scores also capitalise on the transmission of alleles from mother to offspring for SNPs that have a direct effect on own birth weight provided direction of effect is consistent. For the GxE MR analysis, we used parity in females as the environmental variable to stratify our analyses, with the nulliparous group acting as the negative control. Parity was defined by converting the self-reported number of births into a binary indicator. To investigate the possibility of introducing collider bias when conditioning on parity, genetic variants were tested for an association with parity while adjusting for standard covariates (P-value < 0.05/381 = 1.3×10^-4^; Figure S2).

Association analyses were performed between maternal PGS and standardised maternal outcomes (all traits with mean 0 and SD 1) using R (lm) in 171,031 parous and 40,239 nulliparous females, separately. Note the parous group included the 19,553 female participants who had self-reported having an offspring but without reporting the birth weight of their first child. We also performed a combined analysis with both parous and nulliparous females to determine if there was an interaction between the PGS and parity. The presence of an association in parous females, with the absence of an association in nulliparous females provides evidence that some aspect of the intrauterine environment (with maternal genotypes associated with offspring birth weight as a proxy) has causal associations with later life risk for cardiovascular disease. An association in nulliparous females suggests the presence of pleiotropic pathways from the genetic variants to the outcome.

We leveraged the existence of a “no-relevance” negative control (i.e., nulliparous females) to estimate causal effects in the absence of horizontal pleiotropy. For both parous and nulliparous females, causal estimates were obtained via two sample MR. SNP-exposure associations were based on the SNPs used for the maternal PGS for offspring birth weight and the corresponding weights from Hwang et al. (Hwang et al., 2024). SNP-outcome associations were estimated separately in parous and nulliparous females at the same SNPs for each of the cardiometabolic outcomes, following the same approach as described above for the PGS. The Wald ratio was used to obtain causal estimates separately in parous and nulliparous females. Estimates in the nulliparous group provide an estimate of the magnitude of pleiotropy while those in the parous group reflect both potential causal effects and pleiotropy. We subtract the estimated effect in nulliparous females from that estimated in the parous females to obtain a pleiotropy robust causal effect estimate, with standard errors estimated using the variance of the difference (i.e. the sum of the separate variances).

#### Offspring genotype by proxy Mendelian randomization

In the absence of large numbers of genotyped mother-offspring pairs in the UKB, we employed an approach that instead utilises spousal genotypes (i.e. paternal genotypes) to proxy offspring birth weight. To understand why this analysis would be informative, consider Figure 1, which illustrates a credible way in which we can obtain causal estimates of offspring traits on maternal outcomes in the absence of offspring genotype. Because fathers transmit half their alleles to their offspring, it follows that paternal genotype will proxy offspring genotype, which in turn will proxy offspring phenotype. This permits estimation of the causal effect of the offspring trait on the maternal outcome, assuming no horizontal pleiotropy between the paternal genotype and maternal outcome and obviates the requirement of individual-level genotyped mother-offspring pairs. Further, under the assumption of random mating, the use of paternal genetic variants as instruments should also protect MR analyses from confounding from the maternal genome. The offspring genotype by proxy MR approach involves the same three core assumptions as conventional MR analyses, however, under this approach there are additional nuances to these assumptions that need to be borne in mind (Figure S3). We provide a detailed discussion of these assumptions in supplementary material 4.

**Figure 1:**
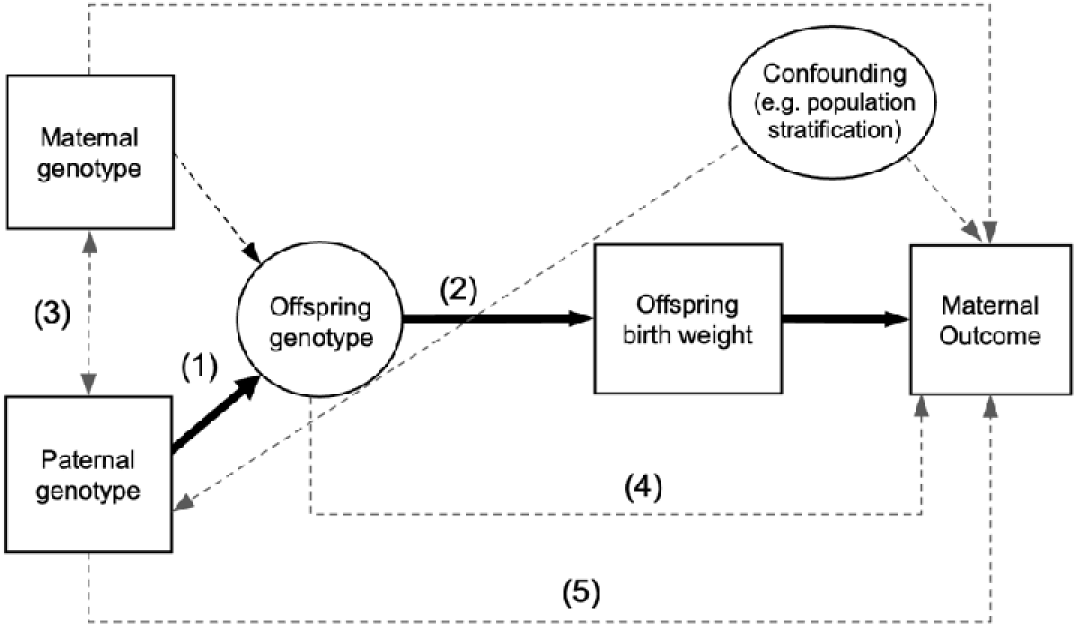
Causal diagrams illustrating the offspring genotype by proxy MR study design to investigate the causal effect of offspring birth weight on maternal cardiometabolic health outcomes. Observed variables are represented by squares, whilst latent unobserved variables are represented by circles. Solid arrows indicate the paths of interest in MR. Dotted arrows indicate secondary paths or assumptions. In the offspring genotype by proxy MR approach, paternal genotype is utilized to proxy offspring genotype (path 1), which in turn proxies offspring phenotype (i.e. offspring birth weight; path 2). Thus, paternal genetic variants can be used as instrumental variables for MR (s indicated by the path in bold). This obviates the need for offspring genotype which is indicated by denoting offspring genotype as a latent unobserved variable. Under random mating, paternal and maternal genotypes should be uncorrelated (path 3), which blocks confounding through the maternal genotype. The design also assumes the absence of pleiotropic paths from the offspring genotype to the maternal outcome (path 4) and direct effects of the paternal genotype on maternal outcome (path 5).

Offspring genotype by proxy MR was performed using the individual level data in the UKB to assess evidence of a causal relationship between offspring birth weight and maternal cardiometabolic risk factors. In the absence of offspring genotype, to proxy the exposure of interest (offspring birth weight) paternal genetic variants for own birth weight were used to proxy the offspring’s genetic variants for own birth weight. PGS for own birth weight were constructed using paternal genetic variants. PGS were constructed from several combinations of SNPs (F1, F2, and F3, as defined above). In the main results we report on the unweighted PGS constructed from SNPs that have an association with own birth weight as instruments for fetal effects on own birth weight. Weighted scores were used to support these analyses, however, owing to sample overlap with the discovery GWAS, these are provided as secondary analyses.

Causal estimates were calculated using two-stage least squares using the IVreg package in R (Fox et al., 2024). In the first stage, offspring birth weight was regressed on the paternal PGS for own birth weight. The first stage regression was then used to estimate genetically predicted offspring birth weight. In the second stage, the maternal outcomes were regressed on genetically predicted offspring birth weight. In addition, we implemented an IVW MR analysis along with weighted median and weighted mode sensitivity analysis using the TwoSampleMR package (Hemani et al., 2018) (https://github.com/MRCIEU/TwoSampleMR). MR Egger regression was not performed as this is less appropriate for MR performed in a single sample (Minelli et al., 2021).

Lastly, we used the two-sample MR approach that allowed for the SNP-exposure association and the SNP-outcome association to be taken from different samples. Here we aimed to improve the accuracy of our causal estimates by leveraging larger sample sizes for the SNP-exposure association. As our SNP-exposure association aimed to capture the effect of offspring genotype on offspring birth weight (and not maternal effects), we used the association between offspring genotype and offspring birth weight conditional on maternal genotype from Hwang et al., (Hwang et al., 2024). The SNP-outcome association was taken from the association of paternal genetic variants for their own birth weight (as a proxy for offspring genetic variants) and the maternal outcomes from the spousal pairs as before. As offspring genotype was used for the SNP-exposure association and paternal genotype for the SNP-outcome association, and given the relationship between offspring genotype and paternal genotype is on average 0.5, we applied a correction of times two to the SNP-outcome association to calculate the causal estimate using a Wald ratio. An additional benefit to the two-sample procedure was the utilisation of spousal pairs that had reported having an offspring, but did not have offspring birth weight recorded, for the estimation of the SNP-outcome association.

## Results

Descriptive statistics for the cohort after quality control are shown in Table S1. Statistical power to detect causal effect estimates are presented in Supplementary Material 3. For the MR GxE approach, we had 80% power to detect a 0.14 increase in standardised outcome per kg increase in offspring birth weight using sample size of 171,000 parous females and an instrumental variable that explained 1.5% of the variance in offspring birth weight (based on an observed effect size of 0.127, variance of offspring birth weight of 0.16 and assuming a two tailed type 1 error rate of 0.05). For the offspring genotype by proxy MR analysis we had 80% power to detect approximate causal effect sizes of 0.48 and 0.40 (i.e. change in our standardised outcomes per kg increase in offspring birth weight) when the proportion of variance explained in offspring birth weight by the paternal PGS was 0.5 and 0.7%, respectively (based on an observed effect size of 0.127 in 45,000 spousal pairs, variance of offspring birth weight of 0.16 and assuming a two tailed type 1 error rate of 0.05; Figure S4).

### Conventional observational epidemiological analyses

The conventional observational analysis, when assessed in 151,478 parous females, identified associations between offspring birth weight and all the maternal cardiometabolic risk factors excluding TC and TG (Figure 2 and Table S4). Increased offspring birth weight was associated with lower maternal SBP, DBP, HDL as well as increased maternal Gl, LDL and BMI. There was no association between offspring birth weight and maternal TC or TG levels. Phenotypic associations were also assessed in the subset of females with a spouse in UKB and found to be largely consistent relative to the entire cohort, suggesting the spousal subset is generally representative of the greater population (Figure 2 and Table S4). Lastly, there was some evidence of an association between offspring birth weight and these paternal phenotypes in fathers (SBP, TG and BMI; Table S4. The magnitude of association was smaller than in mothers for SBP and BMI, with the direction of effect reversed for TG, relative to females.

**Figure 2:**
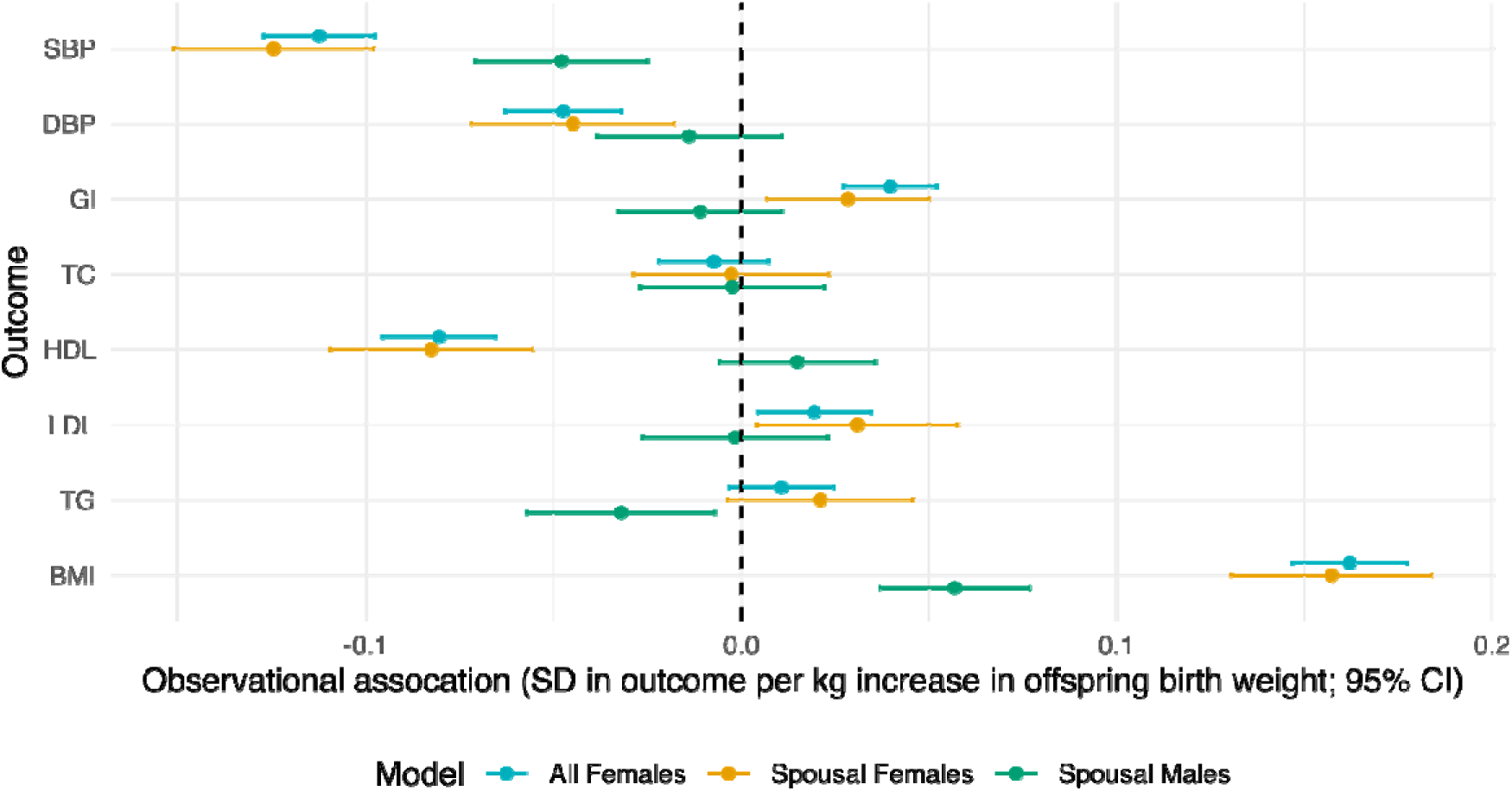
Observational association between offspring birth weight and parental cardiometabolic risk factors in later life estimated using multivariable linear regression. All models were adjusted for potential confounders including age at assessment, assessment centre, genotyping batch, the first 20 genetic PCs, age at first and last birth, number of offspring, smoking status (current, previous or never), alcohol intake frequency (converted to units per month), Townsend deprivation index and years of education. Effect estimates represent the difference in mean standardised outcome for 1 kg higher offspring birth weight and the corresponding 95% CI. Abbreviations: systolic blood pressure, SBP; diastolic blood pressure, DBP; glucose, Gl; total cholesterol, TC; high density lipoprotein cholesterol, HDL; low density lipoprotein cholesterol, LDL; triglycerides, TG; and body mass index, BMI.

### GxE MR

Both the weighted and unweighted maternal PGS for offspring birth weight were positively associated with offspring birth weight in the parous group, explaining between 1.1 and 2.1% of the variation in offspring birth weight (corresponding F-statistic ranging between 1,679 and 3,181; Table S5). For the individual SNPs, 313 out of 381 SNPs were associated with offspring birth weight in the UKB (p<0.05; Table S6). We investigated whether stratifying by parity could introduce collider bias as would be the case if the PGS showed association with this variable. There was no strong statistical evidence of association between parity and the various PGS (P-value > 0.05)(Table S7), nor with the individual SNPs (P-value > 1.3×10^-4^; Table S8).

We found no evidence of differences in the association of the maternal PGS for offspring birth weight and any of the cardiovascular risk factor outcomes between parous and nulliparous females (Figure 3). Specifically, for BMI we observed no evidence of an association of the PGS across all scores. However, for the other traits we observe varying degrees of association, with the PGS putatively pleiotropically influencing both birth weight and the cardiovascular risk factors. This suggests that for these traits there is not a causal effect of offspring birth weight. Instead, the observational association is likely driven by other mechanisms such as genetic pleiotropy. Our results were consistent across the different PGS employed and across both unweighted and weighted strategies (Table S9). Further, we observed no evidence of a formal interaction between the PGS and parity, analytically confirming no difference in the effect of the PGS on the outcome between the two groups (Table S10).We provide pleiotropy robust causal estimates of offspring birth weight on maternal cardiometabolic risk factors in Table S11. Pleiotropy robust causal estimates were calculated by subtracting the estimated causal effect in nulliparous females from parous females.

**Figure 3:**
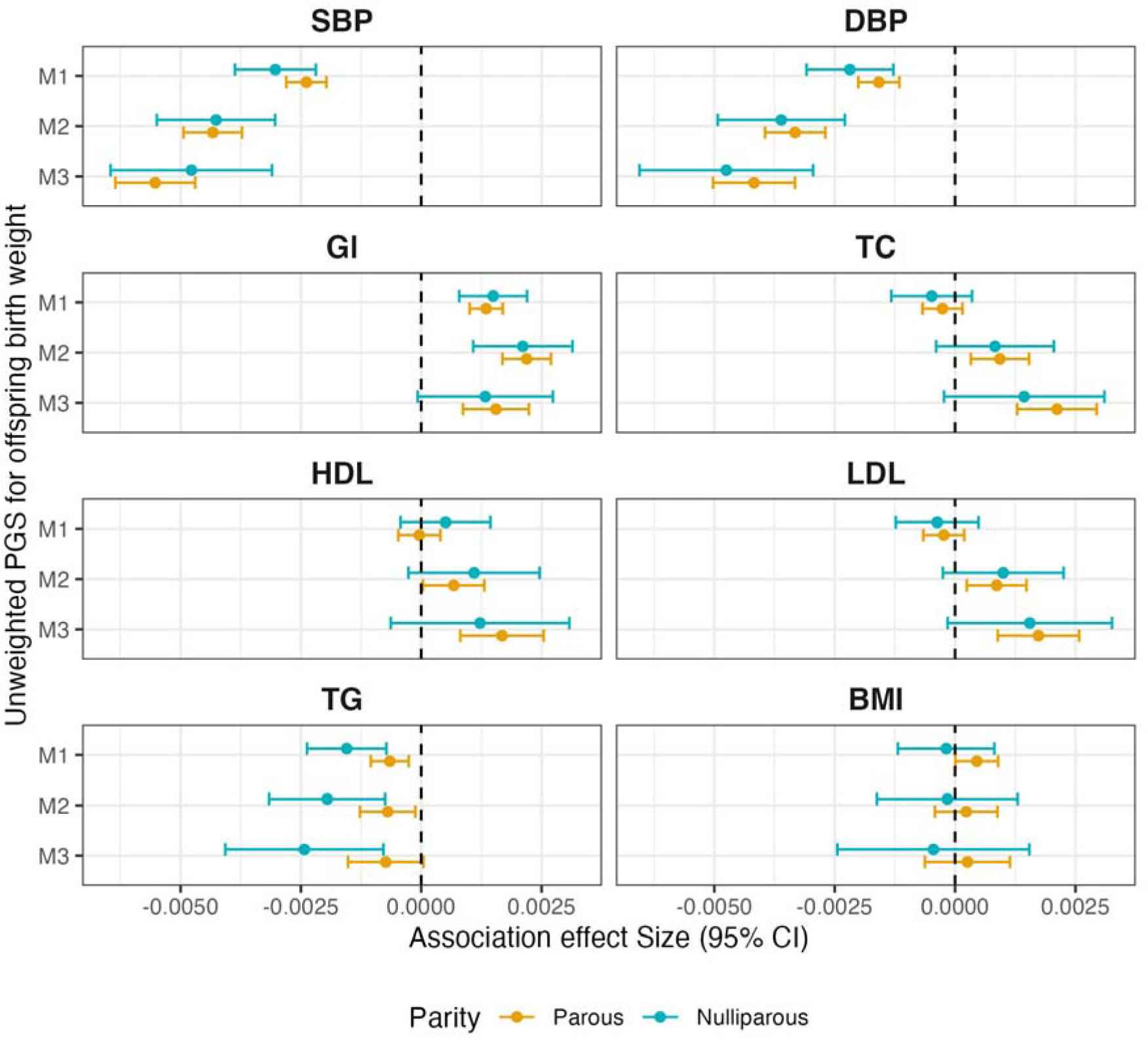
Standardized coefficients for the regression of maternal outcome on maternal PGS in parous (n= 171,031) and nulliparous women (n= 40,239). The regressors were unweighted PGS constructed from SNPs associated with offspring birth weight calculated in females. The outcomes were eight cardiometabolic risk factors measured in females at the UKB assessment centres. Analyses were adjusted for maternal age at assessment, maternal assessment centre, maternal genotyping batch and the first 20 maternal genetic PCs. Parous mothers include those whom did not report offspring birth weight but did report having at least one offspring (n=19,553). Unweighted PGS for offspring birth weight were calculated as the sum of birth weight increasing alleles for 381 birth weight associated SNPs (M1), 176 SNPs that showed significant evidence of a maternal effect (M2) and 94 SNPs that showed evidence of only maternal effects on offspring birth weight (M3). Abbreviations: systolic blood pressure, SBP; diastolic blood pressure, DBP; glucose, Gl; total cholesterol, TC; high density lipoprotein cholesterol, HDL; low density lipoprotein cholesterol, LDL; triglycerides, TG; and body mass index, BMI.

### Offspring genotype by proxy MR

#### Validating assumptions

Our first stage regression demonstrated the three unweighted paternal PGS for own birth weight were positively associated with the offspring birth weight reported by their spouse, with F2 explaining the largest percentage of variance (R^2^ of 0.6% and F-Statistic of 281; Table S12). The proportion of phenotypic variance in offspring birth weight that was explained by the paternal PGS was found to be approximately ¼ of that for own birth weight (2.7% variance explained) which is consistent with quantitative genetics theory. As for the individual SNPs, 305 out of 381 SNPs showed evidence for positive association with offspring birth weight (P-value < 0.05; Table S13). We investigated the assumption of no assortative mating on the exposure and found that the maternal and paternal PGS for own birth weight were uncorrelated (Table S14). To assess violations of the exclusion restriction assumption, we first examined whether birth weight SNPs and the corresponding PGS in fathers were associated with potential confounders, including paternal cardiometabolic risk factors, paternal smoking status, alcohol consumption, deprivation index and years of education. Using a nominal P-value threshold of 0.05, we found evidence of pleiotropy with the paternal PGS for own birth weight, identifying associations with TG, BMI, Gl, deprivation index, SBP, HDL, years of education, smoking status and alcohol consumption (Table S15). At the individual SNP level, we identified 203 pleiotropic SNPs associated with at least one paternal phenotype (Table S16). Sensitivity analyses were performed excluding the 203 pleiotropic SNPs from the paternal PGS used in the offspring genotype by proxy MR as described below.

#### Causal effect estimates

The offspring genotype by proxy MR analyses found no strong evidence for a causal relationship between offspring birth weight and any of the maternal cardiometabolic risk factors in 45,546 spousal pairs when using the paternal PGS for own birth weight as the instrument (Figure 4). This conclusion was consistent across the different instruments employed (Table S17). We performed several MR sensitivity analyses. First, we excluded the potentially pleiotropic SNPs identified above (Table S16) from the PGS. We observed a potential causal effect of offspring birth weight on maternal LDL and TC levels, however these did not pass multiple testing correction (P-value < 0.0125; Table S18). Further, IVW MR weighted median and weighted mode sensitivity approaches found no evidence of a causal effect for any of the maternal outcomes (Table S19). Lastly, when leveraging the two-sample MR approach which utilises SNP-exposure effects from a larger association study, we similarly found no evidence of a causal relationship (Table S20).

**Figure 4:**
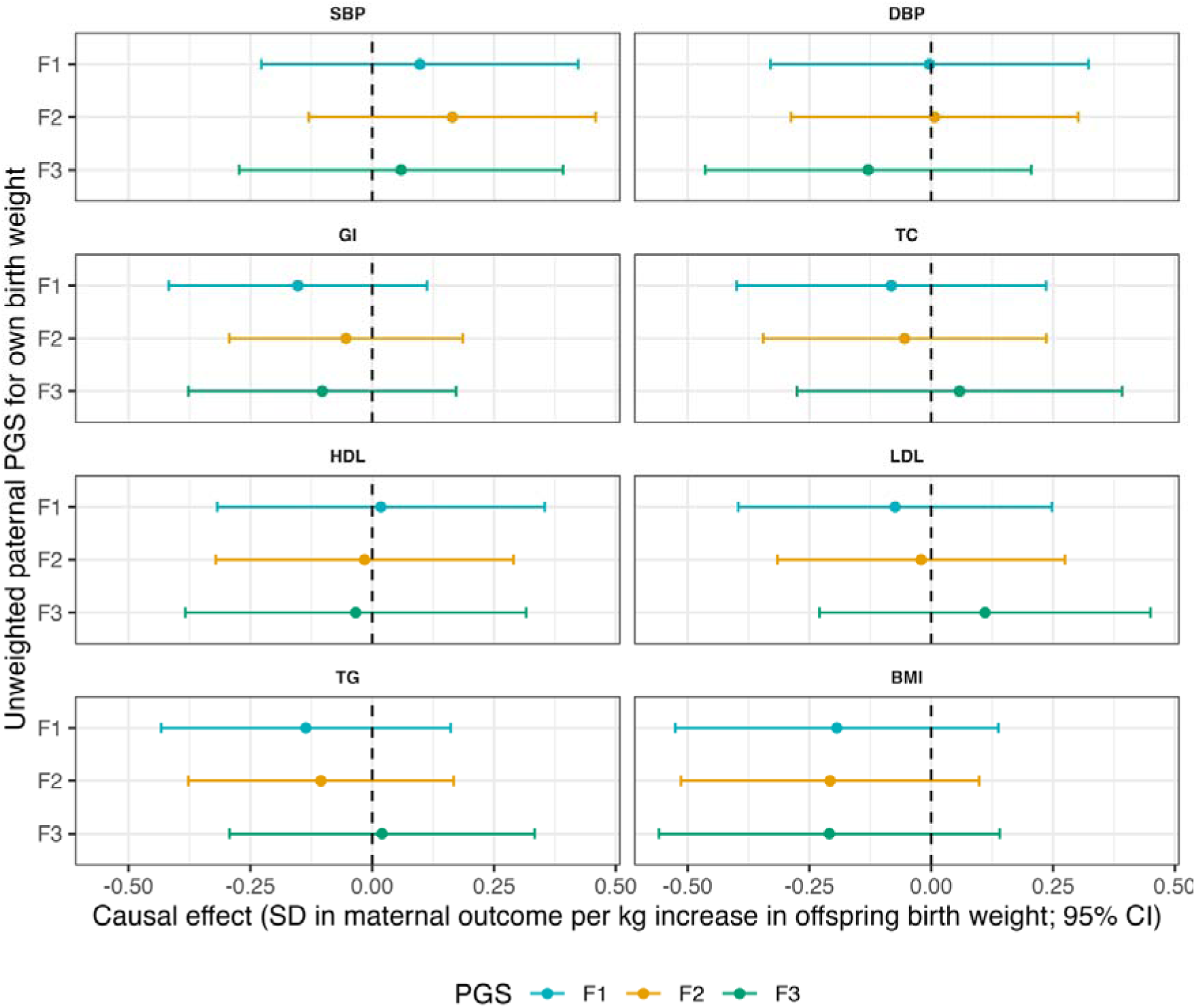
Causal effect estimates of offspring birth weight on maternal cardiometabolic risk factors estimated using the offspring genotype by proxy MR approach calculated in 45,546 parous spousal pairs. The instrumental variables employed were unweighted PGS constructed from SNPs associated with own birth weight calculated in spousal males. The exposure was offspring birth weight reported by spousal females. The outcomes were 8 cardiometabolic risk factors measured in female spouses at the UKB assessment centres. Causal effects were estimating using 2SLS where both stages were adjusted for maternal age at assessment, maternal assessment centre, maternal genotyping batch and the first 20 maternal genetic principal components. Unweighted PGS for own birth weight were calculated as the sum of birth weight increasing alleles for 381 birth weight associated SNPs (F1), 287 SNPs that showed significant evidence of a fetal effect (F2) and 205 SNPs that showed evidence of only fetal effects on own birth weight (F3). Abbreviations: systolic blood pressure, SBP; diastolic blood pressure, DBP; glucose, Gl; total cholesterol, TC; high density lipoprotein cholesterol, HDL; low density lipoprotein cholesterol, LDL; triglycerides, TG; and body mass index, BMI.

## Discussion

In the present study we investigated whether there was a potential causal effect of fetal growth (proxied using offspring birth weight) on maternal cardiometabolic risk in later life. This study was motivated by evidence of fetal influences (including fetal growth) on maternal physiology during pregnancy (Chen et al., 2020; Liu et al., 2014; Petry et al., 2017; Petry et al., 2007; Petry et al., 2016; Petry et al., 2011) and the observational associations between fetal growth and later life maternal cardiovascular disease incidence (Rich-Edwards et al., 2014). We employed two complementary MR approaches which both found no strong evidence for a causal effect of offspring birth weight on maternal cardiometabolic risk. The consistency in results across approaches provides evidence against such a causal effect contributing to previously reported observational associations. Instead, there was strong evidence that maternal genetic variants for offspring birth weight were associated with many cardiometabolic risk factors, even in nulliparous females. This suggests that the phenotypic association between offspring birth weight and maternal later life cardiometabolic risk factors may be partially driven by genetic pleiotropy, with the direction of effects being concordant between the observational analysis and the MR GxE analysis for SBP, DBP, Gl and LDL. Conversely, this was not the case for BMI, with associations for the birth weight PGS centred on the null for both parous and nulliparous women. This suggests the phenotypic association between birth weight and BMI is potentially being driven by factors other than genetic pleiotropy such as unmeasured confounding and/or reverse causation. Indeed, previous MR studies suggest that maternal BMI and maternal blood glucose are both causal for offspring birth weight (Chen et al., 2020; Juliusdottir et al., 2021; Tyrrell et al., 2016).

Along with evidence from previous studies, our results suggest that it is not birth weight itself driving this later life association, but rather, low birth weight may reflect e.g. vascular insufficiency due to reduced or abnormal vascular elasticity and maternal uterine artery flow (Parikh et al., 2021). This in turn impacts maternal spiral artery revascularisation, resulting in an inability to supply adequate oxygen and nutrients to the placenta and fetus. Therefore, having low birth weight offspring likely reflects pre-existing vascular dysfunction in the mother that later predisposes her to cardiovascular disease (including coronary heart disease, stroke, and heart failure) and mortality (Bond et al., 2020; Shaikh et al., 2018; Shaikh et al., 2020). Similarly, at an observational level, it has been reported that many pregnancy complications are preceded by subclinical vascular and metabolic dysfunction (Rich-Edwards et al., 2014). Under this hypothesis, pregnancy may act as a physiological ‘stress’ that unmasks a pre-disposed risk to CVD which also results in adverse pregnancy outcomes i.e. SGA. Consequently, we would not expect to see a causal effect of birth weight itself on later life outcomes as explored here. Further, environmental and behavioural factors (especially smoking) have been suggested to contribute to the inverse observational association between birth weight and CVD risk (Shaikh et al., 2019). For example, Haug et al report that approximately half of the excess cardiovascular risk in females who had SGA offspring could be explained by smoking and that a small proportion may be explained by elevated blood pressure (Haug et al., 2021). Similarly, Shaikh reported a reduction in the risk of CVD mortality when adjusting for maternal diabetes and smoking (Shaikh et al., 2018).

In our epidemiological analyses there was some evidence of an observational association between offspring birth weight and paternal CVD risk factors (BMI and SBP), however these were generally of smaller magnitude than in mothers which is concordant with previous findings (Davey Smith et al., 2007; Shaikh et al., 2018). It has also been shown that offspring birth weight is associated with CVD mortality among aunts and uncles (Shaikh et al., 2020). This supports the hypotheses of environmental and lifestyle factors jointly influencing fetal growth and parental cardiometabolic risk as well as genetic pleiotropy with established CVD risk factors. Pleiotropic associations between own birth weight and future cardiometabolic risk factors have been reported previously (Moen et al., 2020; Warrington et al., 2019). This potentially explains the pleiotropic associations observed in our MR GxE of the maternal PGS for offspring birth weight that includes SNPs with fetal effects (M1 and M2) as some of these SNPs would also influence the mother’s own birth weight and be transmitted to the offspring. However, the associations observed for the maternal PGS for offspring birth weight which only contains SNPs with maternal effects on offspring birth weight (M3), suggests variants with indirect maternal effects on offspring birth weight are also likely to be pleiotropic.

There is conflicting evidence as to whether the relationship between offspring birth weight and maternal cardiovascular mortality extends linearly across the full range of birth weight, with distinct associations observed at the tail ends of the distribution. For example, Haug et al. reported an increased risk of CVD and associated mortality for females with a history of SGA offspring (Haug et al., 2021) whereas Morken et al. observed a relationship between large preterm babies and increased maternal risk of both diabetes and cardiovascular mortality (Morken et al., 2018). This is suggestive of a non-linear relationship which may reflect competing maternal and fetal phenomena (Rich-Edwards et al., 2014). Similarly, findings from Horn et al. found mothers with SGA offspring have different cardiovascular risk profiles later in life compared to those with larger (LGA) offspring (Horn et al., 2020). They suggest SGA offspring reflect adverse maternal vascular health. Conversely, LGA offspring might reflect maternal glucose levels and their effect on fetal growth related to glucose being transferred to the fetus via facilitated diffusion across the placenta and resulting in accelerated fetal growth. The MR models employed here assume a linear effect of the exposure on the outcome and are not equipped to investigate such non-linear relationships. The development of non-linear MR methods is an active field (Burgess et al., 2014; Tian et al., 2023) with current methods yielding some controversial results (Burgess, 2023; Hamilton et al., 2024; Wade et al., 2023). We have also assumed that variation across the distribution of birth weight is informative for inference regarding this causal association. While utilising these SNPs represents a useful starting point for investigation, it may be that small perturbations at the tail end of the normal range are causal, which we do not isolate here. Of note, we have not adjusted birth weight for gestational age as the latter is not reported in the UKB. It is possible that this impacts our ability to make informative inferences. In addition, here we have only investigated causal effect of offspring birth weight for the first offspring and have not taken into account the effect of subsequent pregnancies. However, as fetal growth restriction is thought to be more problematic in a woman’s first pregnancy, with offspring birth weight increasing with birth order (Bohn et al., 2021), this should be sufficient for investigating this hypothesis.

In this study we used two separate approaches to proxy offspring birth weight. In the MR GxE approach, we used PGS (M1, M2 and M3) of SNPs indirectly associated with offspring birth weight, to proxy maternal specific mechanisms underlying fetal growth. While in the offspring genotype by proxy MR approach, the own birth weight PGS (F1, F2 and F3) were intended to serve as a proxy for fetal growth directly through the fetal genome. Importantly, while the proposed mechanism of action for SNPs associated with offspring birth weight between the two approaches differed, under gene-by-environment equivalence, we assume both approaches would identify the same causal effect of offspring birth weight if it were present. We recognise the results of our study may be impacted by winner’s curse owing to sample overlap between our analysis sample and the discovery GWAS used for instrument selection owing to the unavailability of a well-powered independent discovery sample with partitioned maternal and fetal effects. We attempted to alleviate the impact of winner’s curse by employing both weighted and unweighted PGS. While we observed some potential loss of power associated with using an unweighted rather than weighted score, our benchmarking checks demonstrate this was minimal. However, we also recognise that using unweighted scores may disproportionally represent loci with particularly small or large associations.

In the MR GxE analysis, we provide pleiotropy robust causal estimates by subtracting the estimated effect in nulliparous females (which reflects genetic pleiotropy) from that estimated in the parous females (which reflects both potential causal effects and pleiotropy). In doing so, we assume the magnitude of horizontal pleiotropy is the same in parous and nulliparous females. However, it may be that the biological pathways affected by the genetic instruments (other than influencing offspring birth weight) may be different in parous and nulliparous women resulting from pregnancy. For example, maternal SNPs that have indirect effects on offspring birth weight are thought to proxy the latent intrauterine environment (IUE). If it were the case that some aspect of the maternal IUE were causal for later life cardiometabolic risk but birth weight itself was not causal, it follows that genetic variants associated with the maternal IUE would also be associated with maternal cardiometabolic risk. In our MR GxE model this would be captured as pleiotropy (within parous women) as the instrument influences the outcome not through the exposure (birth weight). This would manifest in our model as differing magnitudes of pleiotropy between parous and nulliparous women, potentially biasing the causal estimates.

In the offspring genotype by proxy MR, own birth weight PGS were used to proxy offspring birth weight by transmission of birth weight increasing alleles from the father to the offspring. Given offspring birth weight is only reported by female participants in the UKB, accurate identification of spousal pairs through matching of demographic information in UKB was central to validating the relevance assumption in our application of offspring genotype by proxy MR. While identification of spouse-pairs was performed using similar methods to previous studies (Howe et al., 2019; Robinson et al., 2017; Tenesa et al., 2016), it’s important to note that the spouse-pairs have not been confirmed. We cross-checked with identified trios based on genetic information and find them to be largely concordant suggestive that the spousal pairs derived in this study are genuine (Supplementary Material 1). Even with well-matched spousal data this does not confirm the offspring were shared between the spouses. Our PGS (F1, F2, F3) benchmarking checks demonstrated the paternal PGS for own birth weight explained approximate 0.6% of the phenotypic variance in the offspring birth weight reported by female spouses. This was approximately ¼ of that explained in the fathers own self-reported birth weight (2.7%), which is consistent with quantitative genetic theory and supports the assumption that offspring birth weight reported by female spouses was for an offspring that is genetically related to the male spouse. We note there was a minor deviation in the expected proportion of variance explained by the paternal PGS. While this may be due to a degree of mismatch between spousal pairs or owing to some spousal males not contributing genetic information to the offspring, this may also be due to random variation. Given the mothers report the birth weight of their offspring, incorrect matching in a proportion of the spousal pairs will decrease instrument strength, however as the PGS employed here are strong instruments (Table S12) this is unlikely to introduce bias in the estimation of causal effects.

While there was no strong evidence of an association, as our offspring genotype by proxy MR analyses had limited statistical power, if such an effect of cardiometabolic risk factors exists, it may be small compared to other sources of inter-individual variation. While we employed several PGS combinations in order to increase the number of SNPs used to proxy the exposure and subsequent power, our analyses remained underpowered. In addition, we use the two-sample MR approach with SNP-exposure associations leveraged from the GWAS of own birth weight (n=423,683). With this approach, we still failed to find strong evidence of a causal relationship. Lastly, we note the potential for inclusion of negative controls within this analysis approach by using maternal instruments for offspring birth weight and paternal outcomes (i.e. paternal cardiometabolic risk). This can be used to inform violations of the MR assumptions, however, in the absence of evidence of a causal effect, such an analysis was not included here.

In conclusion, we failed to observe evidence of a causal relationship between offspring birth weight and maternal cardiovascular risk factors later in life. The two approaches we presented have some advantages over traditional observational studies, such as being robust to traditional and certain forms of genetic confounding. They also have advantages over simple MR study designs being robust to certain forms of genetic confounding and permitting informative sensitivity analyses and the analysis of negative controls/no relevance groups. However, both methods suffer from a lack of statistical power suggesting that even larger biobanks/meta-analyses will be required in order for the approaches to attain their full potential.

## Ethics

This project received ethical approval from the Institutional Human Research Ethics committee, University of Queensland (Approval Number 2019002705).

## Data availability

Human genotype and phenotype data from the UK Biobank on which the results of this study were based were accessed with accession ID 53641. The genotype and phenotype data are available upon application to the UKB [http://www.ukbiobank.ac.uk/]. GWAS summary results statistics from the DINGO analysis of birth weight are available from https://doi.org/10.48610/9f69854.

## Acknowledgements

This work was supported by Australian National Health and Medical Research Council (NHMRC) grants (GNT1183074, GNT1157714). D.M.E. and this work were supported by an NHMRC Investigator grant (APP2017942). D.A.L contribution is supported by the UK Medical Research Council (MC_UU_00032/05) and British Heart Foundation (CH/F/20/90003). This research has been conducted using the UK Biobank Resource under Application Number 53641. We would like to thank the UK Biobank participants, interviewers, nurses, and staff that made this study possible.

## Author contributions

A.A.H conceptualized the project, conducted the statistical analysis and drafted the manuscript, C.B.N performed analysis and revised the manuscript, C.F and D.A.L interpreted the data and revised the manuscript, D.M.E conceptualized the project, interpreted the results and revised the manuscript

## Supplementary Tables

Table S1: Demographics for females in the UKB. Demographic information is provided for a) all females and b) spousal females for parity, birth characteristics and conventional demographic factors, with corresponding sample sizes provided. Note that for the purpose of this analysis, spousal females were restricted to those with at least one self-reported offspring.

Table S2: Criteria used to match spousal pairs based on demographic information for UKB participants. Provided are the criteria used for matching, the corresponding phenotype and phenotype code in the UKB.

Table S3: Genetic variants and their association with own and offspring birth weight in Hwang et al. The SEM classification and corresponding effect and alternate alleles, beta, se and pvalues are provided. The inclusion of each genetic variant into the own and offspring birth weight PGS (F1, F2, F3, M1, M2 and M3) are shown.

Table S4: Observational associations between offspring birth weight and cardiometabolic risk factors. Observational associations are shown for a) all females, b) spousal females c) spousal males. All models are adjusted for age at assessment, genotype batch, assessment centre, the first 20 genetic PCs, number of children, female age at first birth, female age at last birth, smoking (current, previous, never), SES (deprivation index), years of education and alcohol intake frequency

Table S5: Offspring birth weight PGS benchmarking against offspring birth weight. Associations between each of the maternal PGS (weighted and unweighted) and offspring birth weight are reported for all females with offspring birth weight recorded in the UKB (n=151,477). Analyses were adjusted for maternal age at assessment centre, genotype batch, assessment centre and the first 20 maternal genetic PCs.

Table S6: Offspring birth weight SNP benchmarking against offspring birth weight. Associations between each of the 381 SNPs and offspring birth weight are reported for all female individuals with offspring birth weight recorded in the UKB (n=151,477). Analyses were adjusted for maternal age at assessment centre, genotype batch, assessment centre and the first 20 maternal genetic PCs.

Table S7: Parity collider test with the PGS for offspring birth weight. Associations between each of the maternal PGS (weighted and unweighted) for offspring birth weight and parity in females are reported for all females in the UKB (n=211,270). Parity was determined for female individuals using their self-reported number of offspring and is defined as a binary indicator (ever versus never). Analyses were adjusted for maternal age at assessment centre, genotype batch, assessment centre and the first 20 maternal genetic PCs.

Table S8: Parity collider test with SNPs for offspring birth weight. Associations between maternal SNPs for offspring birth weight and parity in females are reported for all females in the UKB (n=211,270). Parity was determined for female individuals using their self-reported number of offspring and is defined as a binary indicator (ever versus never). Analyses were adjusted for maternal age at assessment centre, genotype batch, assessment centre and the first 20 maternal genetic PCs.

Table S9: MR GxE associations between each of the maternal PGS for offspring birth weight (weighted and unweighted) and maternal cardiometabolic risk factors. Analysis was stratified by parity in females in the UKB and were adjusted for maternal age at assessment centre, genotype batch, assessment centre and the first 20 maternal genetic PCs.

Table S10: MR GxE associations between each of the maternal PGS for offspring birth weight (weighted and unweighted) and maternal cardiometabolic risk factors with a formal test for an interaction between parity and the maternal PGS. Analyses adjusted for maternal age at assessment centre, genotype batch, assessment centre and the first 20 maternal genetic PCs.

Table S11: Estimated causal effect of offspring birth weight on maternal cardiometabolic risk factors in females in the UKB. Causal effects were estimated using the Wald ratio in the parous and nulliparous subgroups. Causal effect accounting for pleiotropy were obtained by subtracting the causal estimate in nulliparous females from that estimated in parous females.

Table S12: Own birth weight PGS benchmarking against own and offspring birth weight. Own birth weight PGS (weighted and unweighted) were calculated in males only and the following benchmarking performed: a) associations with own birth weight in all males (n=84,828), b) associations with own birth weight in spousal males (n=17,767) and c) associations with offspring birth weight in spousal males (n=45,545). Analyses were adjusted for age at assessment centre, genotype batch, assessment centre and the first 20 genetic PCs.

Table S13: Own birth weight SNP benchmarking against own and offspring birth weight. For each of the 381 SNPs, the following are reported a) associations with own birth weight in males in the UKB b) associations with own birth weight in males in the spousal subset and c) associations with offspring birth weight in males in the spousal subset.

Table S14: Correlation between spouses for n=45,546 spousal pairs. Presented are the Pearsons correlation for a) for own birth weight PGS (weighted and unweighted) and b) phenotypic correlations for cardiometabolic risk factors and potential confounders. Note that spousal pairs were matched in deprivation index as part of demographic matching in the identification of spousal pairs.

Table S15: Association between paternal PGS for own birth wight and potential paternal confounders in male spouses. Potential paternal confounders include paternal cardiometabolic risk factors, paternal smoking status, alcohol consumption, deprivation index and years of education. Age is included as a negative control.

Table S16: Association between paternal SNPs for own birth wight and potential paternal confounders in male spouses. Potential paternal confounders include paternal cardiometabolic risk factors, paternal smoking status, alcohol consumption, deprivation index and years of education. Age is included as a negative control. Presented are trait-SNP combinations with P-value < 0.05.

Table S17: MR causal effect estimates of offspring birth weight on maternal cardiometabolic risk factors from offspring genotype by proxy MR analysis in 45,546 spousal pairs. Causal estimates were calculated using two-stage least squares for each of the own birth wight PGS (weighted and unweighted). Maternal age at assessment was used a negative control. Analyses were adjusted for maternal age at assessment centre, genotype batch, assessment centre and the first 20 maternal genetic PCs.

Table S18: MR causal effect estimates of offspring birth weight on maternal cardiometabolic risk factors from offspring genotype by proxy MR in 45,546 spousal pairs excluding potentially pleiotropic SNPs. Pleiotropic SNPs were identified based on a conservative P-value threshold of 0.05 in Table S16 and subsequently excluded from the paternal PGS employed as the instrumental variable. Causal estimates were calculated using two-stage least squares for each of the own birth wight PGS (weighted and unweighted). Maternal age at assessment was used a negative control. Analyses were adjusted for maternal age at assessment centre, genotype batch, assessment centre and the first 20 maternal genetic PCs.

Table S19: MR sensitivity analyses of offspring birth weight on maternal cardiometabolic risk factors from offspring genotype by proxy MR in 45,546 spousal pairs. Causal estimates were calculated using inverse variance weighted, weighted median and weighted mode. The SNPs utilised were paternal variants and correspond to those employed in each of the own birth weight PGS. Maternal age at assessment was used a negative control. Analyses were adjusted for maternal age at assessment centre, genotype batch, assessment centre and the first 20 maternal genetic PCs.

Table S20: MR causal estimates and sensitivity analyses of offspring birth weight on maternal cardiometabolic risk factors from offspring genotype by proxy MR in 45,546 spousal pairs using the two-sample MR approach. Causal effect estimates are presented for the inverse variance weighted, weighted median and weighted mode approaches. The SNPs utilised correspond to those employed in each of the own birth weight PGS. SNP-exposure associations were obtained from direct effects on own birth weight from Hwang et al., (Hwang et al., 2024). SNP-outcome associations were of paternal genetic variants for their own birth weight and the maternal outcomes in parous spousal pairs. Maternal age at assessment was used a negative control. SNP-outcome analyses were adjusted for maternal age at assessment centre, genotype batch, assessment centre and the first 20 maternal genetic PCs. The causal effect estimates from one-sample MR as calculated in Table S19 are presented for comparison.

## Supplementary Figures

**Figure S1:**
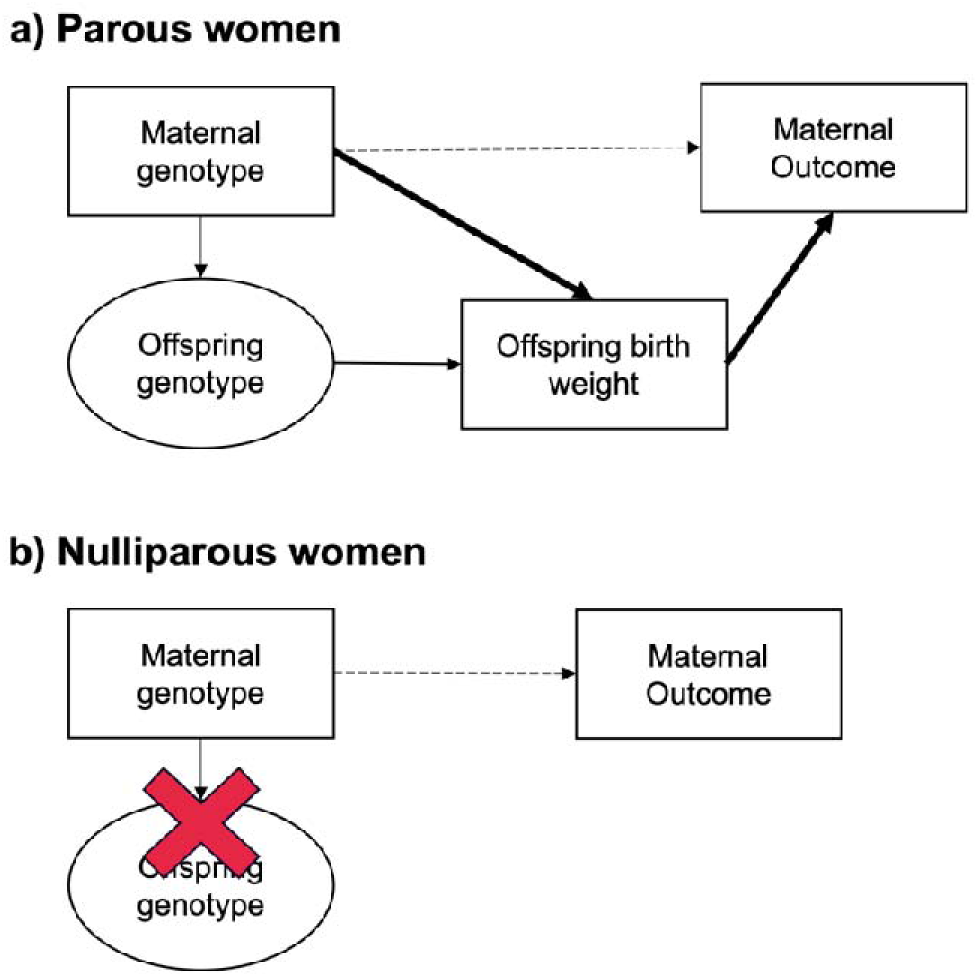
Diagram illustrating the MR GxE study design to investigate a potential causal relationship between offspring birth weight and maternal cardiometabolic risk. Parity was used as the environmental variable on which to stratify the groups, with the nulliparous group acting as a negative control (the “no relevance group”). The association between the maternal genotype and maternal outcome is depicted in (a) parous females and (b) nulliparous females. In parous females, the total effect of maternal genotype on maternal outcome may be comprised of both an effect of the genetic variant through offspring birth weight and an effect through other pathways (e.g. genetic pleiotropy in the maternal genome). In contrast, in nulliparous females there cannot be an effect of the maternal variant through offspring birth weight. Instead, any association between maternal genotype and maternal outcome will likely represent an effect of pleiotropy through the maternal genome. The implication is that the difference in estimated causal effects between the two groups should provide an estimate of the true causal effect of offspring birth weight on maternal outcome free from pleiotropy.

**Figure S2:**
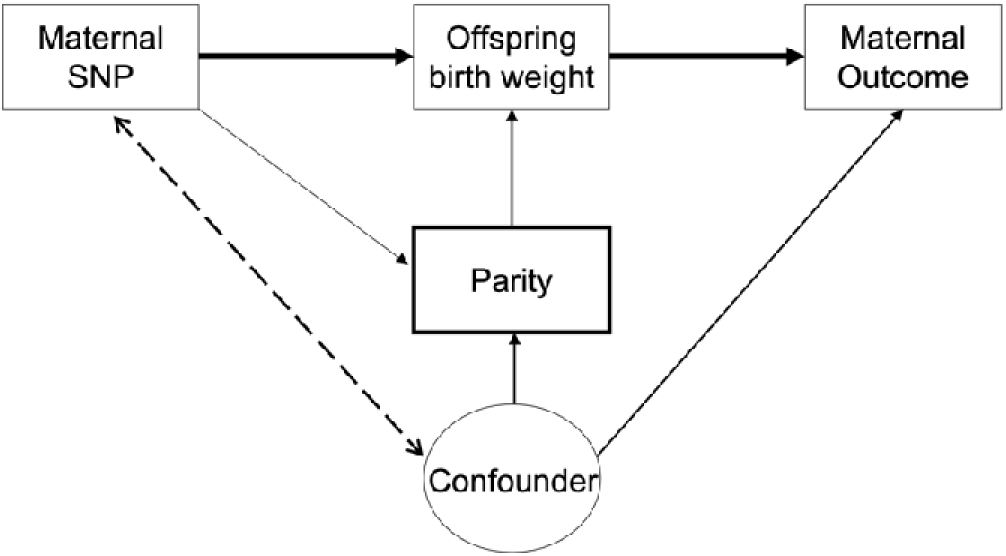
Diagram illustrating a potential case of collider bias in gene-by-environment MR. Collider bias may be introduced when the SNPs used to proxy offspring birth weight (the exposure) are also associated with parity (the variable stratified on). By conditioning (or stratifying by; as demonstrated by the bold box) on the collider (i.e. parity) in the GxE MR framework, a pathway may open between the SNP and the outcome though joint determinants of parity and the outcome. This would induce a spurious association between the IV and the outcome.

**Figure S3:**
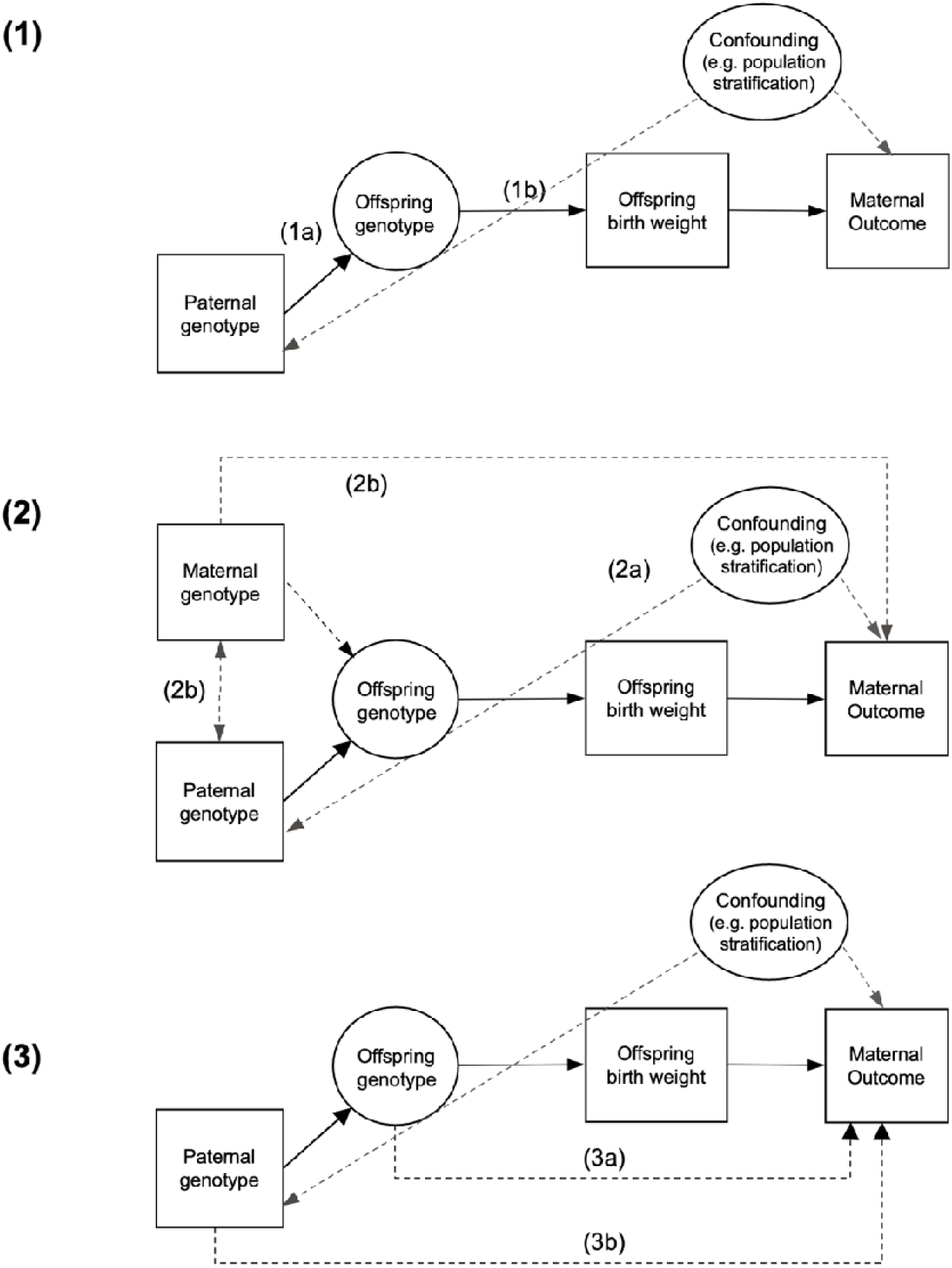
Causal diagram illustrating the offspring genotype by proxy MR approach and the underlying assumptions. Part (1) illustrates the relevance assumption which requires correct matching of spousal pairs using demographic information (1a) and genetic instruments to be associated with the exposure (1b). Part (2) illustrates the independence assumption whereby there are no confounders of the genetic instrument – outcome relationship. This requires genetic instruments to be independent of potential confounders (2a). Further, genetic variants for the exposure in the father should not be associated with genetic variants in the mother (i.e. no assortative mating on the exposure or on traits genetically correlated with the exposure; 2b) as this may create a path from paternal instrument to outcome if those variants (or variants in LD with them) are pleiotropic for the outcome in the mother (i.e. genetic confounding from the maternal genome). Part (3) demonstrates the exclusion restriction assumption which requires no path from paternal instrument to outcome other than through the exposure. This includes the paths from offspring genotype (3a) and paternal genotype (3b) to maternal outcome.

## Supplementary Materials

### Supplementary Material 1: Comparison of criteria used for matching of spousal pairs with trios identified using genetic relatedness

Potential spousal pairs were identified within the UK Biobank using household demographic information (see methods). This included utilisation of the returned spousal file from Tenesa et al. (Tenesa et al., 2016) in addition to further criteria based on household information (Table S2). We performed validation of the spousal pair matching using 1,027 genetically determined parent offspring trios. Whilst two opposite sex individuals having genetically confirmed offspring in the same dataset does not necessarily mean that they are current or former spouses, it is nevertheless likely to be a reasonable assumption in the vast majority of cases. For each of the variables used in the spousal pair definition, the number of matches and mismatches between the genetically determined parents are presented in Table S21. Participants were required to provide identical responses to the household demographic variables, with the exception of household income, where pairs were still matched if their reported income differed by one category or if they had missing income information.

**Table S21.** Distribution of matches and mismatches for household demographic variables used to identify spousal pairs assessed in 1,027 genetically determined mother, father, offspring trios. Variables presented were used in the definition of spousal pairs. Trios were identified based on genetic information with the demographic information reported by the mother and father used for matching.

| **Variables** | **Matches** | **Mismatches** | **Missing Information** |
| --- | --- | --- | --- |
| Tenesa et al., 2015 returned dataset | 764 | 263 | 0 |
| Reported an age difference <10 years | 1026 | 1 | 0 |
| Household relatedness | 921 | 19 | 87 |
| Number in household | 924 | 93 | 10 |
| Age of the parents differed (including age at death)* | 1024 | 0 | 3 |
| Household income | 517 | 163** | 347 |
| Type of accommodation | 1003 | 17 | 7 |
| Townsend deprivation index | 993 | 33 | 1 |
| Own or rent accommodation | 960 | 42 | 25 |
| Number of vehicles in household | 952 | 64 | 11 |
| Number of children (NEB or Number of children fathered) | 973 | 51 | 3 |

Note:
The criteria used to construct the Tenesa et al., 2015 returned dataset included an age difference <10 years, household relatedness, the number in household and differing age of the parents (including age at death).
\* Matches indicate pairs where the age of either parent differed, and mismatches indicate pairs where the ages of both parents were the same. Missing data was classified as a match under the Tenesa definition.
\*\* Mismatches include 152 pairs with 1 category difference.

### Supplementary Material 2: Multiple Testing Correction Threshold

A PCA was performed across 135,888 parous females with complete cardiometabolic phenotypes available to determine the number of PCs that explained >80% of the variance shared between the phenotypes. This value provides an estimate of the effective number of independent traits and can be used for Bonferroni correction. We determine that 4 PCs accounted for 80% of the covariance between outcomes, which suggests an approximate multiple-testing corrected P-value threshold of p < 0.05/4 = 0.0125 for statistical significance. PCA was performed in R using the prcomp function.

### Supplementary Material 3: MR Power calculations

We estimated the statistical power to detect the causal effect of offspring birth weight on maternal cardiometabolic risk factors in our study. For the offspring genotype by proxy MR analysis we used the MR power calculator (https://shiny.cnsgenomics.com/mRnd/) (Brion et al., 2012) to approximate the size of the causal effect that we had 80% statistical power to detect in 45,000 spousal pairs. For all calculations, we assumed Type 1 error rate of 0.05 and estimated the size of the causal effect on the outcome that we had 80% power to detect. We calculated power under several scenarios using the internal estimates of the proportion of variance explained in offspring birth weight by each of the three paternal PGS for own birth weight from the first stage regression analyses (Table S12).We had 80% power to detect approximate causal effect sizes of 0.48 and 0.40 (i.e. change in our standardised outcomes per kg increase in offspring birth weight) when the proportion of variance explained in offspring birth weight by the paternal PGS was 0.5 and 0.7%, respectively (based on an observed effect size of 0.127; Figure S4). For the MR GxE approach, we estimated the size of the causal effect we had 80% power to detect in 171,000 parous females. We calculated power assuming 1.5% of the variance in offspring birth weight was explained by the maternal PGS for offspring birth weight. We had 80% to detect an approximate causal effect size of 0.14 standard deviations in the outcome per kg increase in offspring birth weight.

**Figure S4:**
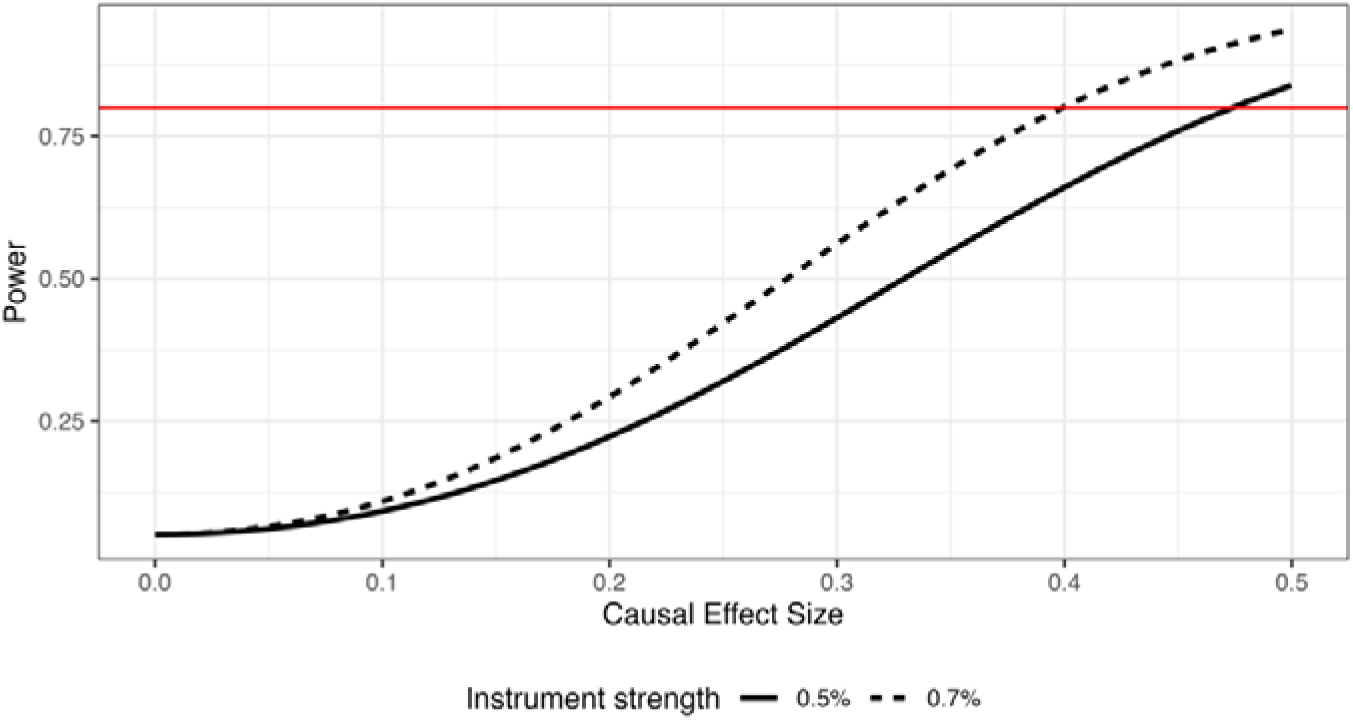
Power calculations for the offspring genotype by proxy MR analyses in 45,000 spousal pairs.

The solid red line represents 80% power, whereas the black lines represent different proportions of variance explained in the exposure by the paternal PGS. Power is calculated using an observed effect size of 0.127 based on the observed phenotypic association between offspring birth weight and maternal BMI (α = 0.05). Figure only presents power calculations for the offspring genotype by proxy MR analyses. Power calculations for the MR GxE analyses are described in text.

### Supplementary Material 4: Assumptions of the offspring genotype by proxy MR approach

The offspring genotype by proxy MR approach involves the same three core assumptions as conventional MR analyses, however, under this approach there are additional nuances to these assumptions that need to be borne in mind.

### Relevance assumption

In the offspring genotype by proxy MR approach, the relevance assumption depends on the accuracy of spousal matching and on the strength of association between the genetic variants and the relevant exposure (Figure S3 part 1). Accuracy of the spousal data requires not only for the spousal pairs to be matched correctly (here based on demographic information) but for offspring birth weight reported by the female participants to be the genetic offspring of the matched male spouse, noting that in in the UKB, only female participants report on the birth weight of their offspring (i.e. male spouses contribute genetic information to the offspring for which female spouses report offspring birth weight). Instrument strength was assessed by examining the association between paternal genetic variants for own birth weight and offspring birth weight reported by female spouses and estimating the proportion of phenotypic variance explained in the exposure by the instruments. We evaluated the strength of the instruments using the proportion of variance in the exposure explained by the instrument (R^2^) and the conditional-F statistic. The reported conditional F-statistic was calculated as F = R^2^ * (n-1) / (1-R^2^). Note that the SNPs used here are known to exert fetal (direct) effects on birth weight, as would be expected when the corresponding alleles are transmitted from father to their offspring. This, however, does not preclude the possible (although in this case, unlikely) existence of paternal (indirect) effects on birth weight at these same loci, which in the one-sample analysis will contribute to the paternal SNP-offspring birth weight association.

### Independence assumption

The independence assumption in MR requires no confounders between the genetic variants and outcomes of interest (Figure S3 part 2). In standard MR, both assortative mating (in previous generations) and population stratification violate this assumption and bias MR estimates (Hartwig et al., 2018). Firstly, we attempt to control for population stratification by adjusting all analyses for the first 20 genome-wide genetic PCs as well as genotyping batch. We consider the implications of assortative mating between spousal pairs on the exposure (Figure S3 path 2b) that may induce a correlation between the genetic instruments in the spouses. If the genetic instruments (or variants in LD with them) are also pleiotropic with the outcome, this could create a path from paternal instrument to maternal outcome. To investigate the possible presence of assortative mating on the exposure, we assessed the correlation in genetic variants between spouses.

### Exclusion restriction assumption

The exclusion restriction assumption stipulates the genetic instrument must only be potentially associated with the outcome through the exposure. In MR, this assumption may be violated by horizontal pleiotropy. For example, offspring genotype may relate to maternal outcome by paths other than through the offspring exposure (Figure S3 path 3a). Because paternal genotype is also related to offspring genotype, then this state of affairs would violate the exclusion restriction assumption. In the offspring genotype by proxy MR design, there is an additional path that also need to be considered (Figure S3 path 3b). First, we note the exclusion restriction assumption will be violated if the exposure of interest, when present in the father, is causal for the maternal outcome. We recognise this path as a limitation in the application of offspring genotype by proxy MR to offspring traits more broadly, however in the case of birth weight we argue that this is unlikely to be problematic (i.e. birth weight in the father is unlikely to be causal for maternal cardiometabolic risk later in life). Another potential violation is if the paternal genetic instrument is pleiotropic for other paternal phenotypes (that are not the exposure) which in turn are causal for the maternal outcome and/or if spousal mating is based on those phenotypes. For example, a proportion of birth weight SNPs have been shown to be pleiotropically associated with BMI (Warrington et al., 2018). Therefore, any association between paternal BMI and maternal BMI, either due to assortative mating or causality, would open a viable path from instrument to outcome other than through the exposure of interest.

We performed sensitivity analyses to investigate these potential pleiotropic effects by first examining whether birth weight SNPs and the corresponding PGS in fathers were associated with potential confounders. This included paternal cardiometabolic risk factors as well as potential paternal confounders such as paternal smoking status, alcohol consumption, deprivation index and years of education. These analyses were performed using linear regression on the subset of male spouses and adjusted for standard covariates (as outlined earlier) in the spousal males. Where associations were identified, additional analyses were repeated in the spousal pairs using the PGS, this time excluding all SNPs associated with any of the above traits (p<0.05/381=1.3×10^-4^).

